# Screening of renal function among a group of physicians working in a hospital who are often self neglected

**DOI:** 10.1101/2021.05.17.21257285

**Authors:** MA Kashem, Ahsan Ullah, Rajat Sanker Roy Biswas

**Affiliations:** Department of Nephrology, Chattagram Maa O Shishu Hospital Medical College, Chittagong, Bangladesh; Department of Medicine, Chattagram Maa O Shishu Hospital Medical College, Chittagong, Bangladesh

**Keywords:** Physicians, Proteinuria, Serum creatinine, Body Mass Index, Modified Diet in Renal Disease formula, Estimated Glomerular Filtration Rate.

## Abstract

**Background:** Prevalence of chronic kidney disease (CKD) has been increasing rapidly worldwide and early screening to detect kidney disease, particularly at its early stages is pivotal to stop its further deterioration. Population based study on detection at early stage of kidney disease and it’s prevalence are scanty in our country, especially among the medical professional personnel. Hence taking advantage of the observance of World Kidney Day 2020, we conducted a screening program for kidney disease organized at the Chattogram Maa O Shishu Hospital (CMOSH) premises among a group of physicians of the hospital.

**Methods:** This was a cross sectional observational study among 67 physicians of different ranks (medical officer to professor), working at our hospital in different discipline. Age, body weight, height, Body Mass Index (BMI), blood pressure were documented, and urinary protein and serum creatinine were measured at a single sitting. Kidney function was estimated by calculating the glomerular filtration rate (GFR) using the Modification of Diet in Renal Disease (MDRD) formula. Kidney function was classified according to estimated GFR (eGFR) and Kidney Disease Outcomes Quality Initiative (K/DOQI) guidelines.

**Results:** A total of 67 working physicians of the hospital were studied. Among them majority subjects (30) were in the age group of 20 to 29 years. Among all 11.9% participants had proteinuria (trace to 1 plus). The distribution of eGFR was symmetrical, with the majority (70.10%) of subjects in the 60 to 89 ml/min category; 11.90% had 30 to 59 ml/min category and only 17.90% of the study population had eGFR > 90 ml/min. An inverse relation between eGFR and age, and a direct relation between eGFR and BMI were observed.

**Conclusion:** Proteinuria, low eGFR levels might be related with future decline in renal function among the studied subjects and so regular health check-up is important to abate the potential epidemic of kidney disease among the health professionals.

## Introduction

Chronic kidney disease (CKD) is a significant burden of diseases worldwide [1]. As it is insidious in onset and has non-specific clinical manifestations, it often goes unrecognized and later presents with severe multisystem complications. The prevalence of CKD worldwide has been estimated to be about 8–16 % [2]. A study in Dhaka, conducted among adults aged 30 years and older, reported a 26% prevalence of CKD [3], while other studies reported 13% prevalence among the urban Dhaka populations aged 15 years and older [4,5]. Another study reported CKD prevalence of at least 7% among the health service providers in Dhaka [6]. A community-based study estimated that 1/3^rd^ of the rural populations were at risk of CKD that largely remained undiagnosed [7].

However, due to the lack of comprehensive national health database, the changing trend of CKD remains unknown among Bangladeshi population. The reported variations in the prevalence of CKD among Bangladeshi populations may be due to cross-sectional study design using populations from different geographic locations within the Dhaka city, using populations of different ages, shorter study period, and modest sample size. The prevalence of CKD varies in different age groups, among males and females, and by socioeconomic conditions and rate of urbanization. Moreover, the prevalence of CKD is also influenced by health risk factors such as diabetes and hypertension

However, reliable statistics on the prevalence of deteriorated renal function among different segments of the community like physicians is rare, with CKD still being an unrecognized health challenge [8]. Among the hospital stuffs and female health workers working in a tertiary care hospital of Chittagong Bangladesh was found that some of them are at CKD stage 3 and all were undiagnosed [9, 10].

Early identification and appropriate nephrological management of patients with mild renal disease has been increasingly recognized as an important opportunity to delay the progression of renal disease [11]. Although some debate exist about how to most effectively evaluate persons with CKD in general population, previous follow-up study showed that participants in Kidney Early Evaluation Program (KEEP) were more likely to seek and receive renal care and experience less mortality and morbidity compared with non-participants [12]. In view of this, renal status in a group of physicians was assessed by measuring proteinuria, serum creatinine and estimating glomerular filtration rate in a kidney screening program, organized on March 14 to coincide with the observance of “World Kidney Day 2020”.

## Method

In the year 2020 for the celebration of World Kidney Day Chattogram Maa O Shishu Hospital, Agrabad, Chittagong, Bangladesh, Department of Nephrology organized a kidney screening program among a group of working physicians of hospital with prior permission from the Institutional Ethical Committee(IEC). A total of 67 physicians of different ranks were included in the study on first come and first serve basis. All subjects were introduced a questionnaire, documenting their age and sex, and personal and family history of health (hypertension, diabetes, and kidney disease) with the assistance of trained volunteers. Physicians having the history of urinary tract infection or CKD were excluded from the study. Each participant underwent weight and height measurements using a calibrated scale and the BMI was calculated as weight (in kilograms) divided by height in square meters and categorized as per the cutoff value for Asian population. Blood pressure (BP) was measured according to the guidelines presented in the JNC VII [13]. Participants were asked to collect a spot urine sample which was then used to detect proteins with urinary strips [14]. Five ml venous blood was collected for measurement of serum creatinine which was done by an enzymatic method on auto analyzer (DADE Behring Limited, USA) in the college laboratory. The glomerular filtration rate (GFR), calculated by using the simplified Modification of Diet in Renal Disease (MDRD) formula.[15] Using these estimated GFRs (eGFRs) and the Kidney Disease Outcomes Quality Initiative (KDOQI) kidney functioning was classified for each subject. According to the KDOQI guidelines to define CKD, we should have at least 3 months history or 3 positive test results. Since we conducted a screening program and tested only once, we used the term “likely CKD” instead of classical CKD. Using these estimated GFRs (eGFRs) and the Kidney Disease Outcomes Quality Initiative (KDOQI) kidney functioning was classified for each subject. According to the KDOQI guidelines to define CKD, we should have at least 3 months history or 3 positive test results.

Data were analyzed by using SPSS 20 (IBM, Armonk, NY, USA). Results are presented as numbers, percentages, mean, standard deviation (SD) and range.

## Results

Demographic and clinical data revealed mean (± SD) age of the physicians was 36.18 ± 12.20 years; 30 physicians were at age group 20-29 years, 15 were at 30-39 years, 9 were at 40-49 years and at > 50 years were 12 physicians. Mean BMI was 26.93 ± 3.76 Kg/m^2^ . mean systolic and diastolic pressure was 122 ± 14 mmHg and 80 ± 8 mmHg, Mean (± SD) of serum creatinine and eGFR (MDRD) was 1.03 ± 0.13 (mg/dl), and 76.03 ± 12.99 (ml/min), respectively.(Table-1). Proteinuria was detected among 8 subjects as of trace (11.9%) as shown in the table 2. The age-wise distribution of eGFR is shown in Table 3. Only 12 (17.90%) participants had eGFR > 90 mL/min, Majority 47 (70.10%) had eGFR 60-89 mL/min; and 08 (11.90%) had eGFR < 60 ml/min(Table 4). There was a tendency of gradual increase in eGFR up to 40 years old group and then a gradual decline with increasing age. (Figure 1) shows the relation between age and eGFR and there was an inverse relation between the two (R^2^=0.217). And again eGFR showed a positive correlation (R^2^= 0.138) with BMI (Figure 2).

**Table 1:**
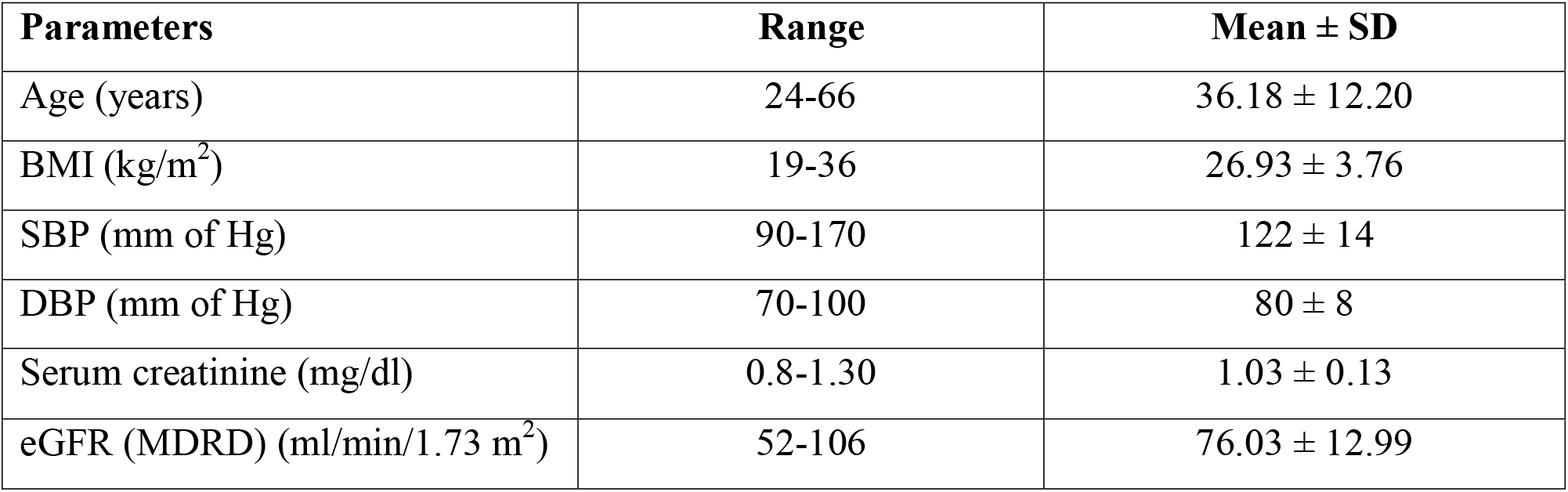
Demographic and clinical data of the studied subject (n=67):

**Table 2:** Status of proteinuria among the study subjects (n=67):

| Proteinuria | Frequency | Percent |
| --- | --- | --- |
| Negative | 59 | 88.1 |
| Trace to positive(≥ 1+) | 08 | 11.9 |
| Total | 101 | 100.0 |

**Table 3:** Age-wise distribution of eGFR calculated by MDRD formula:

| Age group | N | Mean ± SD (ml/min) | Range (ml/min) |
| --- | --- | --- | --- |
| 20- 29 years | 30 | 79.90 ± 14.64 | 60.00-106.00 |
| 30-39 years | 15 | 75.80 ± 10.17 | 60.00-93.00 |
| 40-49 years | 9 | 73.67 ± 9.65 | 60.00-90.00 |
| >50 years | 12 | 68.42 ± 11.17 | 52.00-88.00 |
| Total | 67 | 76.03 ± 12.99 | 52.00-106.00 |

**Table 4:** Stratification of the subjects according to the eGFR (*n* = 67)

| GFR Categories (ml/min/1.73 m<sup>2</sup>) | Number | Percentage |
| --- | --- | --- |
| (I) $\geq 90$ | 12 | 17.90 |
| (II) 60 – 89 | 47 | 70.10 |
| (III) 30- 59 | 08 | 11.90 |
| (IV) 15 – 29 | 00 | 00 |
| (V) <15 | 00 | 00 |

**Figure 1.**
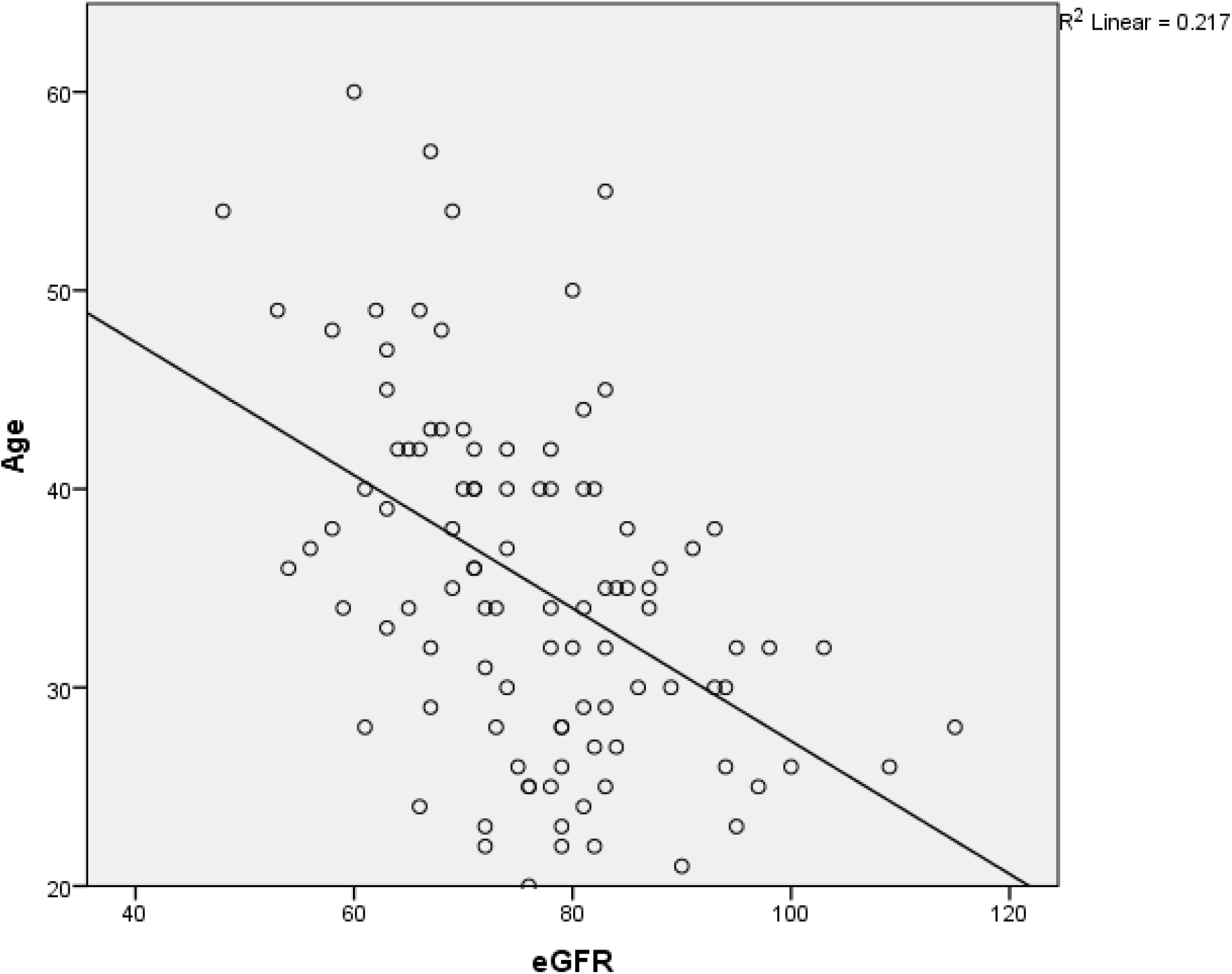
Relation between age and eGFR

**Figure 2:**
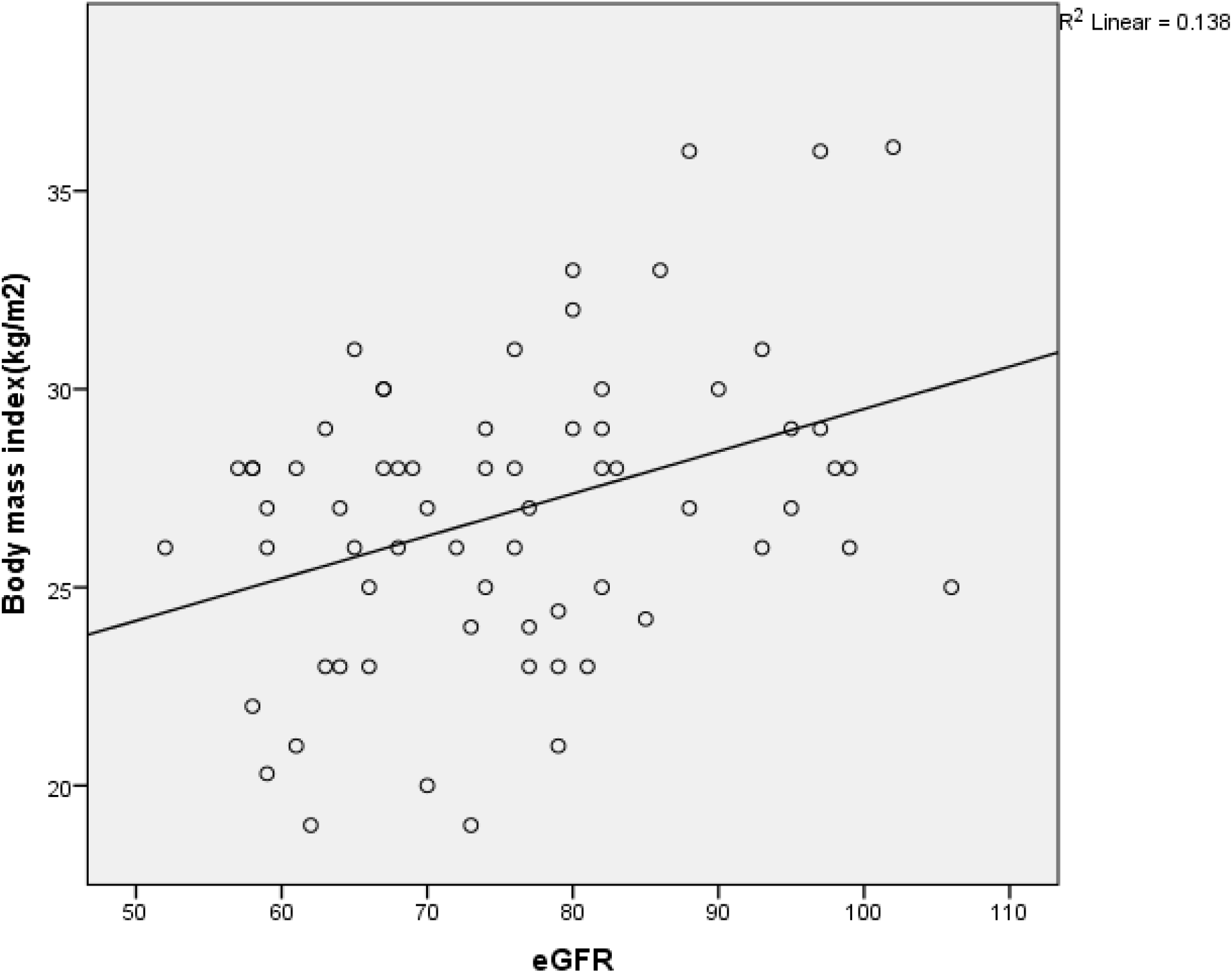
Relationship between the BMI and eGFR

## DISCUSSION

We observed an average eGFR level of 76.03 ± 12.99 mL/min (52.00-106.00 ml/min) in the studied subjects, which is on the lower side of the generally accepted values for normal GFR estimates of 100-110 ml/min [16]. Seventy percent of our subjects had eGFR 60-89 ml/ min category i, e. at CKD stage II and 11.90% subjects had eGFR 30-59 ml/min category i, e. at CKD stage III which is an alarming signal for the working physicians who often neglect themselves in health care. Our previous studies also showed a prevalence of poor or low GFR among hospital stuffs [9] and among the female health workers [10]. As, we do not know the exact average GFR level in our general population hence it is difficult to draw any conclusions on this small data. In general, the risk of developing CKD is higher with increasing age and in our study 18% participants were in the age of more than 50 years. Hence early and regular health screening is very vital to avoid the risk of kidney diseases development in the later life. Recently there has been increasing emphasis on appropriate treatment by timely referral of subjects with early renal disease to the concern specialist [17].

As per the K/DOQI definition of CKD, we observed 11.90% of likely CKD in the stage 3; 70.10% in stage 2 and only 18.0% in stage 1 i.e. eGFR > 90 ml/min. in our study. The apparently lower level of eGFR results in problems of staging due to fixed cut-offs value in the K/DOQI classification of CKD stages. Actually, it is not known if the K/DOQI classification can be applied to our population or not. Whether the lower level of GFR in the population indicates a greater susceptibility to developing CKD or proportionately lower cut-off values at each stage are required is not known. Besides, another contributing factor may be related to the issue of measurement methodology and calibration of serum creatinine [18]. On the whole, the burden of CKD in this small population was much higher compared to the available data in previous study [19] The difference may be partly due to the fact that the earlier studies did not depend on GFR/eGFR but on serum creatinine and clinical judgment only. Moreover single measurement of eGFR in our study might overestimate the prevalence of CKD in our population which demands to follow the subjects by re-testing the same individual more frequently.

We observed a linear negative correlation between age and eGFR. The age-wise distribution of GFR indicated that the peak was in the age group of 30–39 years. The proportion of variation in GFR explained by age was 32.0%. Age is a determinant of renal function and GFR is believed to decline by 1 □ ml/min/1.73 m^2^ per year after the age of 30 □ years in healthy persons [20] Our data is in agreement with other reports based on cross-sectional data, but slightly more than the decline based on longitudinal data done in home and abroad [9,21].

The correspondence between eGFR and serum creatinine values among the various stages of CKD led to the interesting observation that serum creatinine levels were not elevated even in CKD stage III, though the GFR indicated derangement or presence of disease. Only in CKD stages IV and V could elevated levels of serum creatinine be seen. Thus, if we depend on serum creatinine alone, there is a possibility of missing the diagnosis of the disease when it is in its earlier stages and similar views have also been reported previously [22]. Only, one-time measurements of serum creatinine, and a lack of calibration of the measurement of serum creatinine are some of the limitations of the study.

We found proteinuria among 8 subjects as trace. Although majority subjects had proteinuria in trace in amount, the significance of this finding cannot be over looked, as proteinuria signifies underlying renal disease and hence it demand further evaluation. In the contrary, the urine sample was not a first morning sample which does bring up question of false-positive and – negative result. Moreover, there is also the potential of transient proteinuria on a one time dipstick test. We also observed a prevalence of hypertension (SBP ≥ 140 and DBP ≥90 mm of Hg) among 13% participants. However, more community-based studies with longitudinal follow-up are mandatory to address the issues.

The main limitation of this screening program is that the participants were tested only once and some false positive results could have added to the prevalence CKD in some of the subjects with transient changes in parameters like proteinuria, eGFR etc. In contrary, the advantage of this cross-sectional study is that here recruited only working physicians of a hospital who can be followed-up in the future to see the effect of aging on GFR and to detect the false positive cases. Future works should continue on new biomarkers for kidney disease identification that can be used in screening and research on new pharmacologic agents for more effective protection from kidney disease damage or progression.

## Conclusion

Since, the single measurement of proteinuria, hypertension and eGFR in our study might overestimate its prevalence and a follow-up study is mandatory to disclose the exact scenario prevailing in the community and hence to abate the potential epidemic of CKD.

## Data Availability

Data available

## REFERENCES

1. Murray CJ, Lopez AD. Global mortality, disability, and the contribution of risk factors: global burden of disease study. Lancet. 1997;349(9063):1436–1442.

2. Jha V, Garcia-Garcia G, Iseki K, Li Z, Naicker S, Plattner B, et al. Chronic kidney disease: global dimension and perspectives. Lancet. 2013;382:260–272.

3. Das SK, Afsana SM, Elahi SB, Chisti MJ, Das J, Mamun AA, et al. Renal insufficiency among urban populations in Bangladesh: A decade of laboratory-based observations. 2019; PLoS ONE 14(4): e0214568.

4. Anand S, Khanam MA, Saquib J, Saquib N, Ahmed T, Alam DS, et al. High prevalence of chronic kidney disease in a community survey of urban Bangladeshis: a cross-sectional study. Globalization and health 2014;10: 1.

5. Huda MN, Alam KS, Harun Ur R. Prevalence of chronic kidney disease and its association with risk factors in disadvantageous population. Int J Nephrol 2012: 267329. pmid:22848823

6. Fatema K, Abedin Z, Mansur A, Rahman F, Khatun T, Sumi N, et al. Screening for chronic kidney diseases among an adult population. Saudi Journal of Kidney Diseases and Transplantation 2013; 24: 534–41.

7. Das S, Dutta PK Chronic kidney disease prevalence among health care providers in Bangladesh. Mymensingh Med J 2010 19: 415–421.

8. Hasan MJ, Kashem MA, Rahman MH, Qudduhush R, Rahman M, Sharmeen A, et al. Prevalence of Chronic Kidney Disease (CKD) and Identification of Associated Risk Factors among Rural Population by Mass Screening. Community Based Medical Journal 2013;1: 20–26.

9. Kashem MA, Biswas RS, Mamun SM. Kidney disease screening among a low-income group of hospital staffs who have less opportunity. J Sci Soc 2020;47:13–6.

10. Kashem MA, Biswas RS, Jewel KH. Kidney disease screening in a group of female health personnel: Who are often missing? J Integrate Nephrol Androl 2019 (in press).

11. Jungers P. Screening for renal insufficiency: Is it worth while? Is it feasible? Nephrol Dial Transplant 1999;14: 2082–2084.

12. Kurella Tamura M, Li S, Chen SC et al. Educational programs improve the preparation for dialysis and survival of patients with chronic kidney disease. Kidney Int 2014;85(3): 686–692.

13. Chobanian AV, Bakris GL, Black HR, et al. The Seventh Report of the Joint National Committee on Prevention, Detection, Evaluation, and Treatment of High Blood Pressure: The JNC 7 report. JAMA 2003;289: 2560–72.

14. Yamagata K, Iseki K, Nitta K, et al. Chronic kidney disease perspectives in Japan and the importance of urinalysis screening. Clin Exp Nephrol 2008;12:1–8.

15. Levey AS, Greene T, Kusek JW, Beck GJ. Simplified equation to predict glomerular filtration rate from serum creatinine. J Am Soc Nephrol 2000;11:828(A).

16. Delanaye P, chaeffnerE Ebert N, Cavalier E, Mariat-Jean-Maie C, Moranne O. Normal reference values for glomerular filtration rate: what do we really know? Nephrol Dial Transplatation. 2112, 27 (7): 2664–2672.

17. Gungers P. Screening for renal insuffiency : is it worth while? Is it feasible. Nephrol Dial Transplant 1999, 14;2082–2084

18. Coresh J, Astor BC, Greene T, Knoyan G, Levey AS. Prevalence of chronic kidney disease and decreased kidney function in the adult US population: Third National Health and Nutrition Examination Survey. Am J Kidney Dis 2003;41:1–12.

19. Agarwal SK, Dash SC, Irshad M, Raju S, Singh R, Pandey RM. Prevalence of chronic renal failure in adults in Delhi, India. Nephrol Dial Transplant 2005;20:1638–42

20. Lindeman RD, Tobin JD, Shock NW. Association between blood pressure and rate of decline in renal function with age. Kidney Int 1984;26:861–8.

21. Kawamoto R, Kohara K, Tabara Y, Miki T, Ohtsuka N, Kusunoki T, Yorimitsu N. An association between body mass index and estimated glomerular filtration rate. Hypertens Res 2008; 31 (8):1559–64.

22. Lindeman RD, Tobin J, Shock NW. Longitudinal studies on the rate of decline in renal function with age. J Am Geriatr Soc 1985; 33:278–85.

